# Cross-cultural communication in a women’s health service: A mixed-methods evaluation

**DOI:** 10.1101/2025.09.03.25333025

**Authors:** Georgia Griffin, Safa Ghannam, Yun Su Lin, Mee Mee Zaw, Mahzad Mahdavisharif, Siu Ng, Thi Kim Anh Nguyen, Natalie Williams, Nasrin Javid, Jane Warland, Zoe Bradfield

**Affiliations:** Curtin School of Nursing, Curtin University, Bentley, Australia; Women and Newborn Health Service, Subiaco, Australia; School or Nursing and Midwifery, College of Health Medicine and Wellbeing, University of Newcastle, Gosford, Australia; University of Adelaide, Adelaide, South Australia, Australia

**Keywords:** Communication, Communication Barriers, Cultural Safety, Interpreter Services, Limited English Proficiency, Medical Interpreting, Women’s Health Services

## Abstract

**Background:** Effective communication is critical to safety and quality in healthcare. Women with limited English proficiency face cultural and linguistic barriers to quality communication in anglophone settings, perpetuating poor health outcomes and health inequity. Women’s perspectives are essential to inform future evidence-based strategies. This study aimed to evaluate and explore women’s experiences of cross-cultural communication within a women’s health service.

**Methods:** Women receiving maternity or gynaecology care at a women’s health service in Western Australia were eligible to participate if they had accessed interpretation support in Arabic, Burmese, Farsi, Mandarin or Vietnamese. A total 68 women completed a cross-sectional survey; 15 women participated in a semi-structured telephone interview. Quantitative and qualitative data underwent descriptive statistical and multilingual reflexive thematic analyses respectively.

**Results:** Participants reported diverse language preferences and skills, most frequently identifying Mandarin, Arabic and Vietnamese as their preferred language. At their most recent visit, most recalled having an in-person interpreter present speaking their preferred language. However, less than half of participants were provided with written health information resources in their preferred language. Three themes, *the role of interpretation*, *navigating the health service with uncertainty*, and *a foundation of trust*, explore facets of women’s experiences, linked by an overarching theme, *Health is too important not to understand*.

**Conclusions:** This study highlights the need for proactive service-wide approaches to cross-cultural communication encompassing but not exclusive to, clinical encounters, offering insight into key systemic gaps. Improved integration of interpretation services, language-concordant information resources and language-concordant support for service navigation are recommended.

**Plain language abstract:** Effective communication is critical for safety and quality in healthcare. Barriers to cross-cultural communication perpetuate health inequities. This study found that participants experienced inconsistent access to and quality of professional interpretation, provision of language-concordant health information resources was inadequate, and language support was limited to clinical interactions, impeding service navigation. Proactive strategies are recommended to improve service-wide cross-cultural communication not only in clinical encounters, but in women’s broader interactions with health services.

## Introduction

Communication is critical to the provision of safe and effective healthcare(1). This is recognised internationally (2,3) and in national policy in many high resource settings. In Australia, this is reflected in the National Safety and Quality Health Service Standards against which Australian health services are accredited(4). Effective communication is acknowledged as integral to promote patient safety, and to enable shared decision-making and informed consent, requiring service-wide systems and strategies to be in place(4).

These National Standards are supported by the Australian Charter of Healthcare Rights, emphasising consumer rights to open and honest communication(5), and a specific user guide recommending tailored communication strategies for healthcare consumers from migrant and refugee backgrounds(6). Health service policies operationalise national standards and guide implementation of such strategies at a local level, such as the provision of interpretation services.

Healthcare settings are increasingly culturally and linguistically diverse(7, 8). Language barriers are recognised as impeding effective communication and thereby consumer safety in healthcare(9). In addition to language, culture informs how both people and organisations communicate, including verbal communication, print materials, online and audio-visual information mediums (10). The term cross-cultural communication, sometimes used interchangeably with culturally responsive communication or transcultural communication, highlights the cultural lenses that influence communication between people and organisations with different cultures(7). The importance of this was highlighted in a rapid review of culturally responsive communication in Australian healthcare settings(7). The authors found that culturally responsive communication can result in improved consumer adherence to recommended treatment, and in improved consumer retention and understanding of health information. Culturally responsive communication was also found to improve consumer and healthcare provider (HCP) satisfaction, and consumer health overall(7).

Cultural and linguistic barriers may intersect to inhibit effective communication when consumers are not proficient in the language in which care is delivered. Reports from Australia show that for women with limited English proficiency (LEP) in healthcare settings, language barriers may intersect with gendered cultural values and limited health literacy, resulting in disengagement and dissatisfaction with care, reduced involvement in decision-making and autonomy, and poorer health outcomes overall (11–13). One notable Australian study (13) reported that, compared to their Australian-born counterparts, women from non-English speaking backgrounds were more likely to be dissatisfied with perinatal care, feel uninformed and that their preferences were not taken into consideration. Despite the introduction of state-wide maternity reforms during the eight-year study period, little improvement in non-English speaking women’s satisfaction with care was identified. The authors theorised these findings reflected a persistent lack of tailored services and inadequate use of interpretation services(13).

Professional interpretation services can be provided within healthcare settings to support linguistically diverse consumers. An international integrative review reported that professional interpretation in hospital settings enhances communication between HCPs, improves patient comprehension and reduces interpretation errors, when compared to relying on family members or untrained bilingual staff(14). The authors also reported variable impacts on length of inpatient stay, lower readmission rates, improved informed consent processes and follow-up care, and positive patient satisfaction associated with professional interpretation. Similarly a systematic review of international research into communication interventions to support people with LEP in healthcare reported patient satisfaction with professional interpretation, identifying in-person interpretation followed by videoconferencing as patients’ preferred interpretation modalities(15). Prior research into communication interventions identified in this review focused largely on interpretation, with little investigation into communication more broadly.

When communication is not effective, poor health outcomes and health inequity are perpetuated(15). To improve health outcomes, insight into cross-cultural communication experiences of people with LEP in anglophone hospital settings is needed(16). This study aimed to explore and evaluate the cross-cultural communication experiences of women who communicate with interpreter assistance at a women’s health service.

## Methods

### Methodology

An exploratory mixed-methods design underpinned by a pragmatic paradigm was selected to enable the exploration of a complex phenomenon, cross-cultural communication(17). Pragmatism allowed for a rigorous flexible design maintaining a focus on answering the research question(18). In Phase One, a descriptive cross-sectional approach was employed to collect data benchmarking women’s experiences and satisfaction with cross-cultural communication at the service. In Phase Two, a qualitative approach employing reflexive thematic analysis was selected, well-suited to an inductive exploration of participant experiences(19). Culturally responsive aspects of project design have been reported elsewhere(20, 21).

### Research setting

This study was conducted in Western Australia, the largest state in Australia, in the geographically isolated, multicultural capital city of Perth(22, 23). As of the 2021 Australian Census, 32.2% of Western Australians were born outside of Australia(23). A language or languages other than English were spoken at home by 18.7% of Western Australians(23).

The study was set within a women’s health service providing maternity and gynaecological care, comprising two hospital sites, one birth centre and a community homebirth program. The number of women who access interpretation or translation services are not routinely monitored or reported by the service. Interpretation and translation of written materials are provided through hospital-employed and contracted organisations external to the service on an as-needed basis. At the time of study design, the most frequent languages for which interpreters were booked at the study site were Arabic, Burmese, Farsi, Mandarin and Vietnamese. With the exception of Farsi, this reflects common language groups in Western Australia with high proportions of LEP(23).

### Participant recruitment and sampling

Women were eligible to participate if they had accessed gynaecological or maternity care at the health service within two years of participating in the study and had utilised hospital-booked interpreter services in one of five languages: Arabic, Burmese, Farsi, Mandarin or Vietnamese.

Non-probability convenience sampling was employed, appropriate to research with traditionally hard-to-reach populations(24). For Phase One, as a cross-sectional descriptive survey was employed, no statistical calculations of sample size were required or conducted(25). A sample size of 15 women was deemed sufficient to capture variety of experience while allowing for depth of reflexive thematic analysis in Phase Two(26).

Multilingual flyers were disseminated throughout waiting areas at the health service, community-based organisations and social media groups. Flyers contained a QR code linked to a language-concordant (translated) participant information sheet and online survey portal hosted on REDCap(27).

Participants could choose to participate in the survey anonymously online or by telephone. Women who expressed interest in a telephone survey were contacted by a language-concordant bicultural research assistant. Women could also choose to participate in a telephone interview. Some women chose to only participate in an interview. All recruitment and data collection materials were made available in Arabic, Burmese, Farsi, English, Mandarin and Vietnamese.

### Data collection

The survey tool and interview guide were developed by a team of clinician researchers and bicultural bilingual research assistants to address the research aim. The survey tool comprised 29 items including demographic items and both single-response and multiple-choice items capturing participant communication preferences and experiences. Three open-ended items captured insights into women’s preferences. The interview guide comprised eight questions to allow further exploration of participants’ communication experiences. Allowing the order of questions to be flexible and prompts such as ‘can you say more about that’ facilitated exploration of participant responses. The English language survey tool and interview guide are available in supplementary material.

All interviews were conducted by a language-concordant bicultural research assistant. Some participants spoke both English and their primary language during the interview, at their own preference. Eight interviews were conducted with an English-speaking-only co-interviewer with clinical experience. This decision was made on review of preliminary interview data because the English-speaking-only interviewer’s clinical experience facilitated further exploration of participants’ experiences. These interviews were conducted in the participant’s primary language and English. Data were collected from July 2023 to August 2024.

## Data analysis

### Quantitative analysis

Multilingual survey responses planned for quantitative analysis, such as country of birth, were translated by bicultural research assistants within the team. Data were then entered in English into SPSS Statistics version 29(28) for descriptive statistical analysis. Frequencies and valid percentages were calculated, excluding missing responses, appropriate to the study aim.

### Qualitative analysis

Audio-recorded interviews were transcribed verbatim in the language spoken during the interview. Qualitative interview and survey data were professionally translated to English. Reflexive thematic analysis was conducted following the six phases set out by Braun and Clarke(19) on 153 responses to open-ended survey items from 62 women, and interview transcripts and field notes from 15 interviews. These phases included familiarisation with the dataset; coding; initial theme generation; theme development and review; refinement, definition and naming of themes; and writing up. For analysis, survey participants were allocated a participant number, such as P1. Interview participants could choose to suggest a pseudonym or have a pseudonym allocated to them.

Reflexive thematic analysis was conducted by the six team members who had conducted the interviews, including GG, SG, YSL, MM and TKAN. An adapted approach to reflexive thematic analysis of a multilingual dataset was undertaken, in which the investigators moved between the source language and English-language translations. Positionality statements of those team members involved in reflexive thematic analysis are available in supplementary material. Once preliminary themes were described, interview participants were invited to participate in synthesised member checking to involve them in the interpretation of the data, thereby enhancing the trustworthiness of the findings(29). Four participants, all Arabic-speaking, chose to participate in this process, following which themes and subthemes were refined.

### Ethical considerations

Careful consideration was given to language, culture, literacy and power differentials between the research team and potential participants throughout the design and conduct of this study. On the treatment of language, recruitment and data collection materials were made available in all study languages. A multilingual multicultural research team, inclusive of interpreter, consumer and clinician experience, was brought together to enable a culturally responsive approach to the research.

## Results

In total, 68 women completed the survey, 14 by telephone and 54 online. A total of 15 women participated in a semi-structured telephone interview ranging in length from four minutes to 71 minutes (mean 34 minutes). Thirteen participants consented to being audio-recorded. Notes were taken for the two participants who did not consent to being recorded and also during telephone surveys and interviews to facilitate later interpretation of the data.

### Participant characteristics

#### Survey participants

Of 68 participants who completed the survey, 47 (69.1%) attended the study setting for maternity and 21 (30.9%) for gynaecology care. Participants had lived in Australia between 8 months and 25 years (mean 7.7 years, median 6 years), and were born in one of 15 countries, most frequently China (n=21, 31.3%) or Vietnam (n=11, 16.4%). When asked to describe their ethnicity, participants nominated 28 different responses, reflecting the diversity of participants within the five language groups. Both ethnicities and languages are reported as described by participants. Common ethnicities included Han (n=14, 21.9%) and Vietnamese or Viet Nam (n=6, 9.4%). Most participants reported no religion (n=24, 37.5%) or being Muslim (n=22, 34.4%). When asked their primary language, 14 languages were reported, most frequently Mandarin (n=19, 28.4%), Arabic (n=13, 19.4%) or Vietnamese (n=11, 16.4%). Five participants (7.5%) reported two primary languages. Most participants reported being able to read in their primary language well or very well (n=64, 97.0%) and write in their primary language well or very well (n=62, 95.4%).

Twenty-two participants (34.4%) reported being able to read in English well or very well and 21 participants (32.3%) reported being able to write in English well or very well. For further details see Table 1.

**Table 1.**
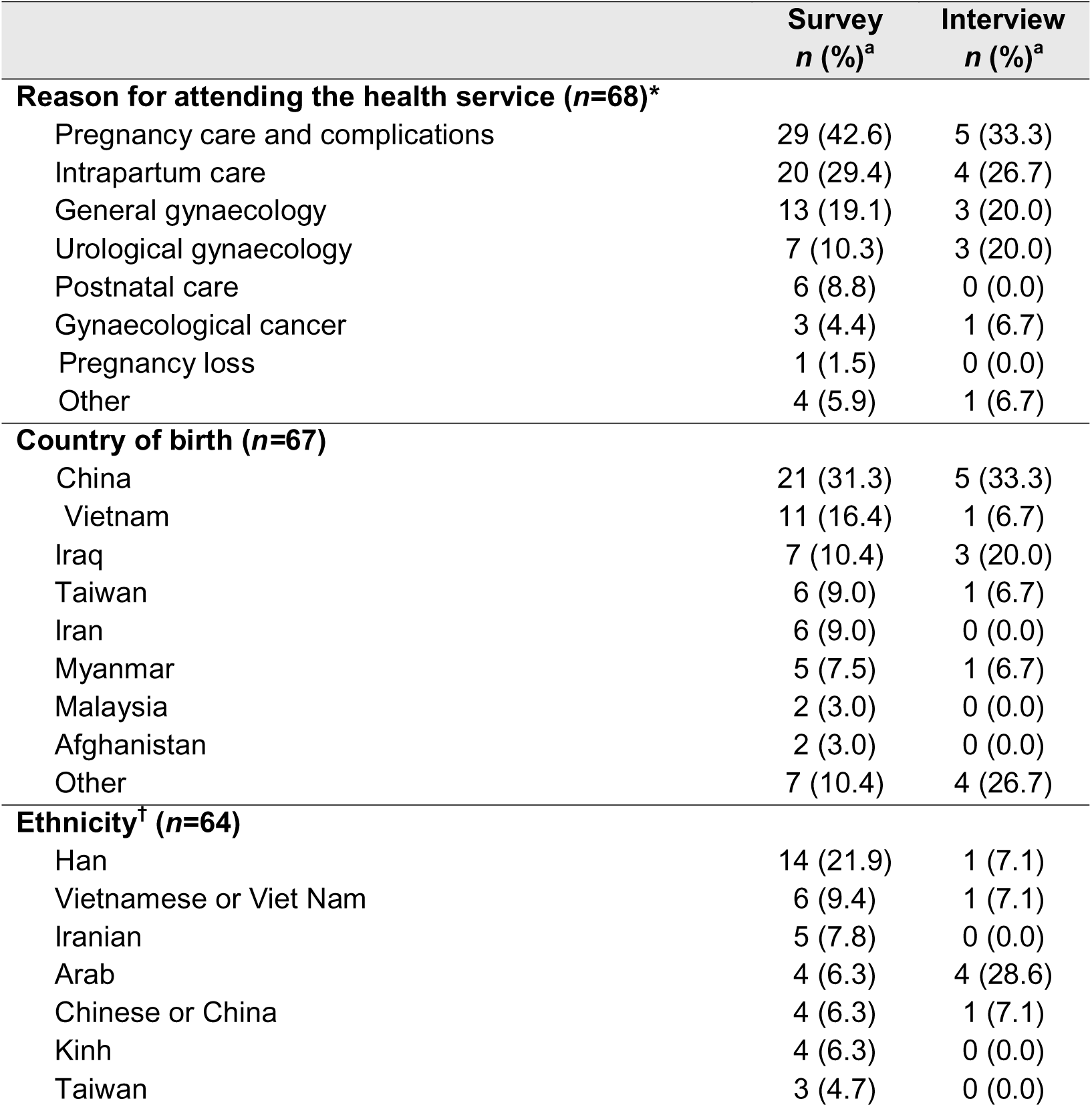
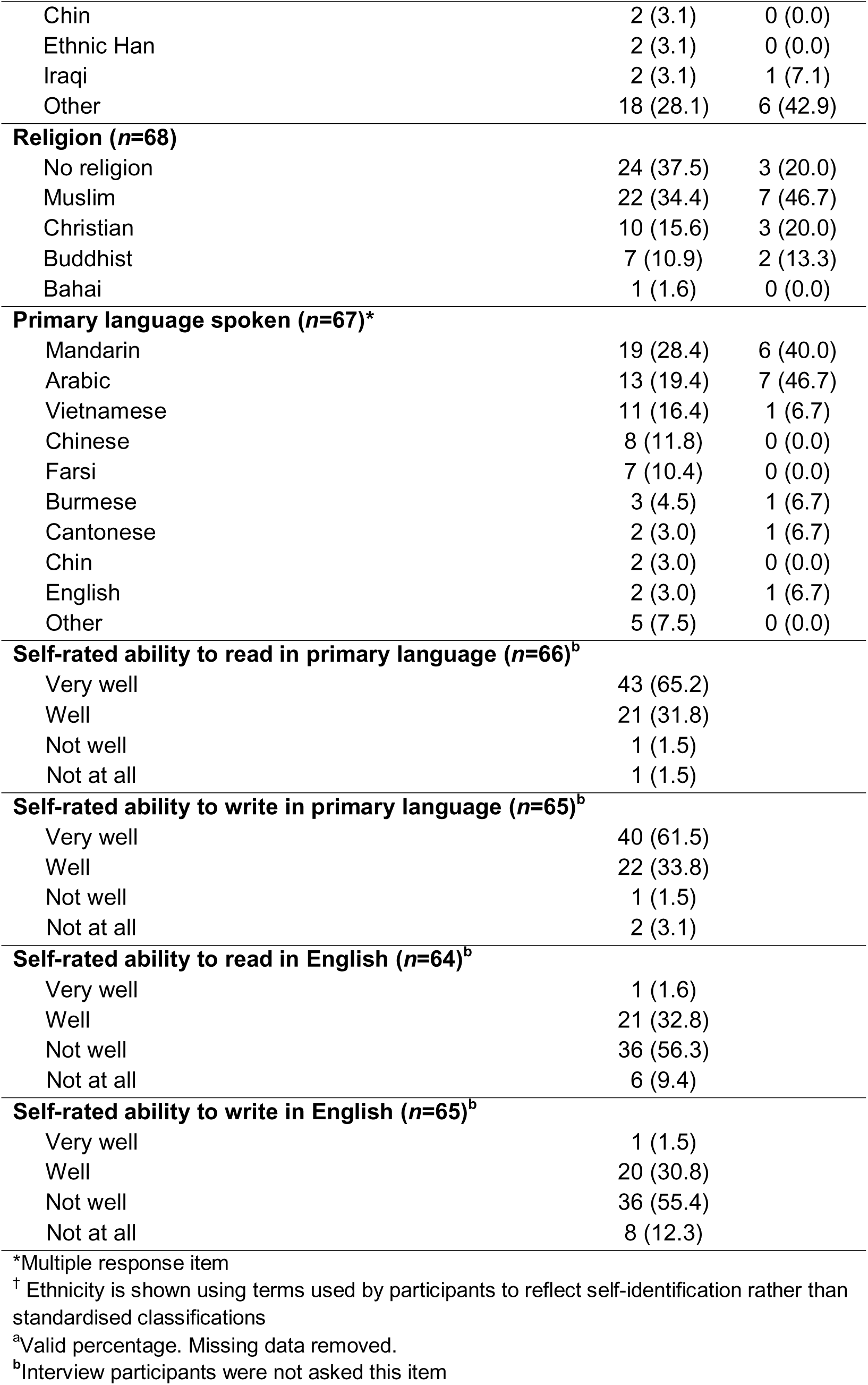
Participant characteristics.

|  | **Survey**<br>**n (%)<sup>a</sup>** | **Interview**<br>**n (%)<sup>a</sup>** |
| --- | --- | --- |
| **Reason for attending the health service (n=68)\*** |  |  |
| Pregnancy care and complications | 29 (42.6) | 5 (33.3) |
| Intrapartum care | 20 (29.4) | 4 (26.7) |
| General gynaecology | 13 (19.1) | 3 (20.0) |
| Urological gynaecology | 7 (10.3) | 3 (20.0) |
| Postnatal care | 6 (8.8) | 0 (0.0) |
| Gynaecological cancer | 3 (4.4) | 1 (6.7) |
| Pregnancy loss | 1 (1.5) | 0 (0.0) |
| Other | 4 (5.9) | 1 (6.7) |
| **Country of birth (n=67)** |  |  |
| China | 21 (31.3) | 5 (33.3) |
| Vietnam | 11 (16.4) | 1 (6.7) |
| Iraq | 7 (10.4) | 3 (20.0) |
| Taiwan | 6 (9.0) | 1 (6.7) |
| Iran | 6 (9.0) | 0 (0.0) |
| Myanmar | 5 (7.5) | 1 (6.7) |
| Malaysia | 2 (3.0) | 0 (0.0) |
| Afghanistan | 2 (3.0) | 0 (0.0) |
| Other | 7 (10.4) | 4 (26.7) |
| **Ethnicity<sup>†</sup> (n=64)** |  |  |
| Han | 14 (21.9) | 1 (7.1) |
| Vietnamese or Viet Nam | 6 (9.4) | 1 (7.1) |
| Iranian | 5 (7.8) | 0 (0.0) |
| Arab | 4 (6.3) | 4 (28.6) |
| Chinese or China | 4 (6.3) | 1 (7.1) |
| Kinh | 4 (6.3) | 0 (0.0) |
| Taiwan | 3 (4.7) | 0 (0.0) |
| Chin | 2 (3.1) | 0 (0.0) |
| Ethnic Han | 2 (3.1) | 0 (0.0) |
| Iraqi | 2 (3.1) | 1 (7.1) |
| Other | 18 (28.1) | 6 (42.9) |
| **Religion (n=68)** |  |  |
| No religion | 24 (37.5) | 3 (20.0) |
| Muslim | 22 (34.4) | 7 (46.7) |
| Christian | 10 (15.6) | 3 (20.0) |
| Buddhist | 7 (10.9) | 2 (13.3) |
| Bahai | 1 (1.6) | 0 (0.0) |
| **Primary language spoken (n=67)\*** |  |  |
| Mandarin | 19 (28.4) | 6 (40.0) |
| Arabic | 13 (19.4) | 7 (46.7) |
| Vietnamese | 11 (16.4) | 1 (6.7) |
| Chinese | 8 (11.8) | 0 (0.0) |
| Farsi | 7 (10.4) | 0 (0.0) |
| Burmese | 3 (4.5) | 1 (6.7) |
| Cantonese | 2 (3.0) | 1 (6.7) |
| Chin | 2 (3.0) | 0 (0.0) |
| English | 2 (3.0) | 1 (6.7) |
| Other | 5 (7.5) | 0 (0.0) |
| **Self-rated ability to read in primary language (n=66)<sup>b</sup>** |  |  |
| Very well | 43 (65.2) |  |
| Well | 21 (31.8) |  |
| Not well | 1 (1.5) |  |
| Not at all | 1 (1.5) |  |
| **Self-rated ability to write in primary language (n=65)<sup>b</sup>** |  |  |
| Very well | 40 (61.5) |  |
| Well | 22 (33.8) |  |
| Not well | 1 (1.5) |  |
| Not at all | 2 (3.1) |  |
| **Self-rated ability to read in English (n=64)<sup>b</sup>** |  |  |
| Very well | 1 (1.6) |  |
| Well | 21 (32.8) |  |
| Not well | 36 (56.3) |  |
| Not at all | 6 (9.4) |  |
| **Self-rated ability to write in English (n=65)<sup>b</sup>** |  |  |
| Very well | 1 (1.5) |  |
| Well | 20 (30.8) |  |
| Not well | 36 (55.4) |  |
| Not at all | 8 (12.3) |  |

\*Multiple response item
<sup>†</sup> Ethnicity is shown using terms used by participants to reflect self-identification rather than standardised classifications
<sup>a</sup>Valid percentage. Missing data removed.
<sup>b</sup>Interview participants were not asked this item

#### Interview participants

From 15 participants who participated in an interview, 8 (53.3%) attended the study setting for maternity care and 7 (46.7%) for gynaecological care. They were aged 30 to 76 years (mean 46 years, median 38 years). They had lived in Australia one to 26 years (mean 9.1 years, median 6 years) and were born in one of eight countries, most frequently China (n=5, 33.3%) or Iraq (n=3, 20.0%). Interview participants reported eight ethnicities, most frequently Arab (n=4, 28.6%). Most described themselves as Muslim (n=7, 46.7%). When asked their primary language, six languages were reported, most frequently Arabic (n=7, 46.7%) and Mandarin (n=6, 40.0%). Both survey and interview participant characteristics are presented in Table 1.

### Evaluation of verbal communication

At their most recent visit to the health service, interpretation services were most frequently provided in Mandarin (n=22, 34.4%), Arabic (n=13, 20.3%) and Vietnamese (n=10, 15.6%). Equal proportions (48.5% v 48.5%) of participants recalled that interpretation services were provided because they had requested or because staff had offered. Most (n=67, 98.5%) reported that the interpreter spoke their preferred language. Most (n=67, 98.5%) also reported that they felt confident to request for an interpreter at the study site. Interpretation modality was most frequently in-person (n=62, 91.2%), aligning with participant preferences. Most participants selected in-person interpretation as their preferred modality (n=57, 93.4%). Twenty-two participants (32.4%) recalled interpretation being provided by telephone despite only two participants (3.3%) identifying telephone interpretation as their preferred modality. Table 2 shows participant perspectives of verbal communication facilitated by an interpreter.

**Table 2.**
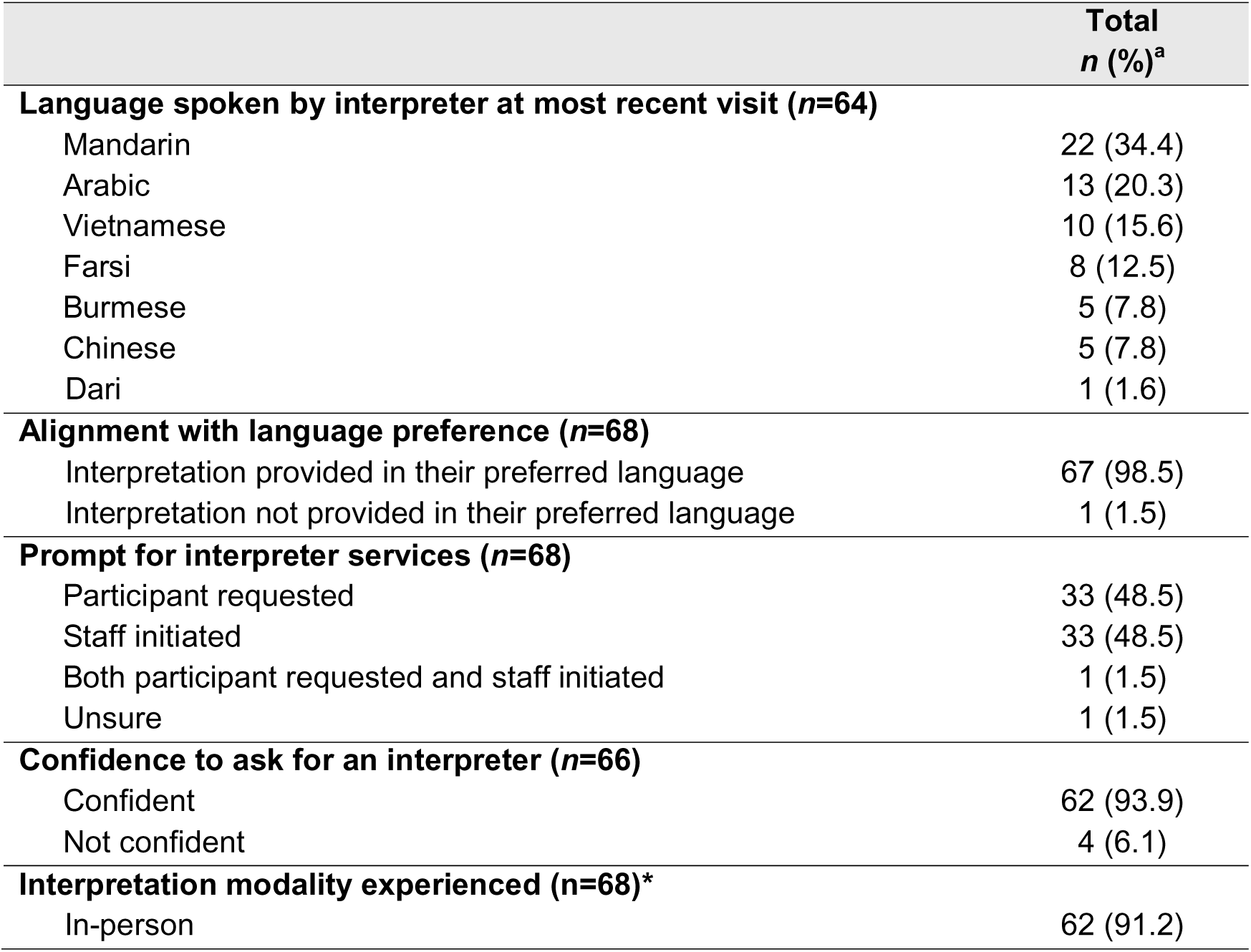
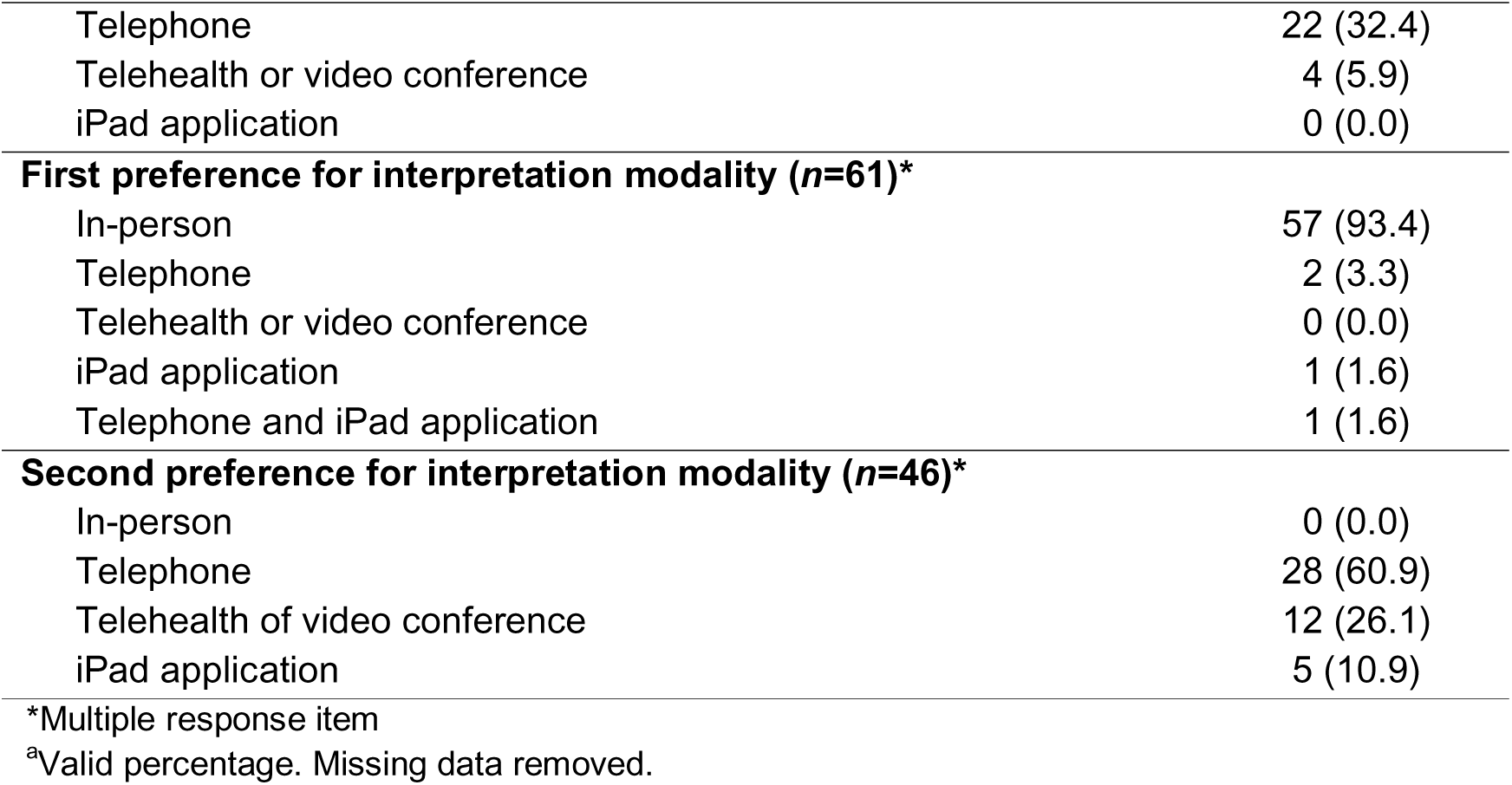
Perspectives of verbal communication facilitated by an interpreter.

|  | **Total**<br>**n (%)<sup>a</sup>** |
| --- | --- |
| **Language spoken by interpreter at most recent visit (n=64)** |  |
| Mandarin | 22 (34.4) |
| Arabic | 13 (20.3) |
| Vietnamese | 10 (15.6) |
| Farsi | 8 (12.5) |
| Burmese | 5 (7.8) |
| Chinese | 5 (7.8) |
| Dari | 1 (1.6) |
| **Alignment with language preference (n=68)** |  |
| Interpretation provided in their preferred language | 67 (98.5) |
| Interpretation not provided in their preferred language | 1 (1.5) |
| **Prompt for interpreter services (n=68)** |  |
| Participant requested | 33 (48.5) |
| Staff initiated | 33 (48.5) |
| Both participant requested and staff initiated | 1 (1.5) |
| Unsure | 1 (1.5) |
| **Confidence to ask for an interpreter (n=66)** |  |
| Confident | 62 (93.9) |
| Not confident | 4 (6.1) |
| **Interpretation modality experienced (n=68)\*** |  |
| In-person | 62 (91.2) |
| Telephone | 22 (32.4) |
| Telehealth or video conference | 4 (5.9) |
| iPad application | 0 (0.0) |
| **First preference for interpretation modality (n=61)\*** |  |
| In-person | 57 (93.4) |
| Telephone | 2 (3.3) |
| Telehealth or video conference | 0 (0.0) |
| iPad application | 1 (1.6) |
| Telephone and iPad application | 1 (1.6) |
| **Second preference for interpretation modality (n=46)\*** |  |
| In-person | 0 (0.0) |
| Telephone | 28 (60.9) |
| Telehealth of video conference | 12 (26.1) |
| iPad application | 5 (10.9) |

\*Multiple response item
<sup>a</sup>Valid percentage. Missing data removed.

Most participants reported feeling comfortable communicating with the assistance of an interpreter (n=66, 97.1%). Factors that promoted comfort were most frequently a friendly HCP (n=44, 65.7%), a friendly interpreter (n=44, 65.7%) and a female HCP (n=34, 50.7%). Factors varied by language group and multiple responses could be selected. For example, for those in the Arabic language group, a female HCP was the most common factor (n=10, 76.9%) whereas in the Mandarin language group, the most common factor was a friendly HCP (n=24, 82.8%). Factors promoting comfort are presented in Figure 1 by survey language.

**Figure 1.**
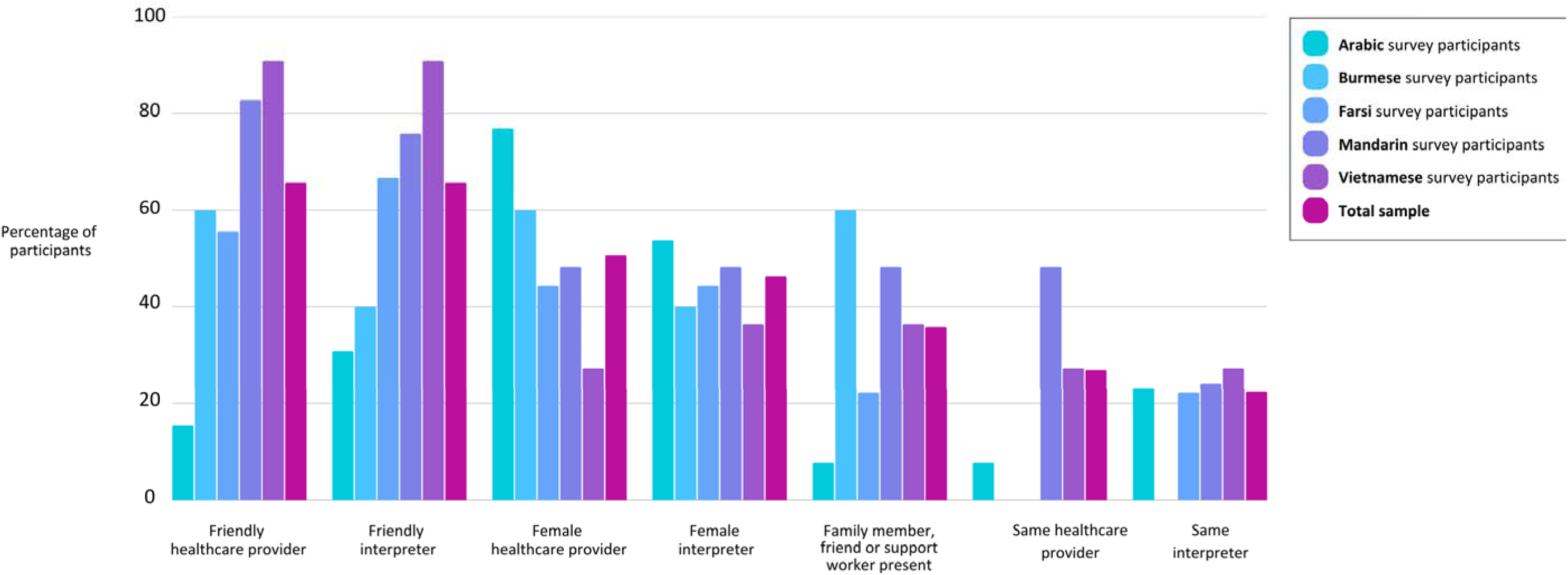
Factors promoting comfort when communicating with a healthcare provider by survey language

### Evaluation of written communication

Most participants reported feeling confident to ask for written health information (n=58, 86.6%). However, while 70.6% of participants (n=48) were offered written information in English, only 44.8% of participants (n=30) preferred to receive written information in English. Most participants (n=52, 77.6%) preferred written information to be provided in the language spoken by the interpreter present. Only 42.6% of participants (n=29) were offered written information in that language. Table 3 shows participants’ perspectives of written communication.

**Table 3.** Perspectives of written communication.

|  | **Total**<br>**n (%)<sup>a</sup>** |
| --- | --- |
| **Confidence to ask for written information (n=67)** |  |
| Yes, confident | 58 (86.6) |
| Not confident | 9 (13.4) |
| **Language of written information offered (n=68)\*** |  |
| English | 48 (70.6) |
| Language spoken by the interpreter | 29 (42.6) |
| In a different language or languages | 1 (1.5) |
| No information offered | 3 (4.4) |
| Not sure | 3 (4.4) |
| **Language preference for written information (n=67)\*** |  |
| English | 30 (44.8) |
| In the language spoken by the interpreter | 52 (77.6) |
| In a different language or languages | 2 (3.0) |
| No written information wanted | 4 (6.0) |

\*Multiple response item
<sup>a</sup>Valid percentage. Missing data removed.

### Exploration of women’s cross-cultural communication experiences

From qualitative interview and survey data, one overarching theme, three themes and eight subthemes were constructed. An overarching theme, *Health is too important not to understand*, illustrated how women’s experiences were connected by an emphasis that good health and effective care were their priority. Communication occurred in the context of feeling unwell, worried, fearful or tired, which made effective communication more difficult and good health outcomes all the more important. A thematic map is presented in Figure 2.

**Figure 2.**
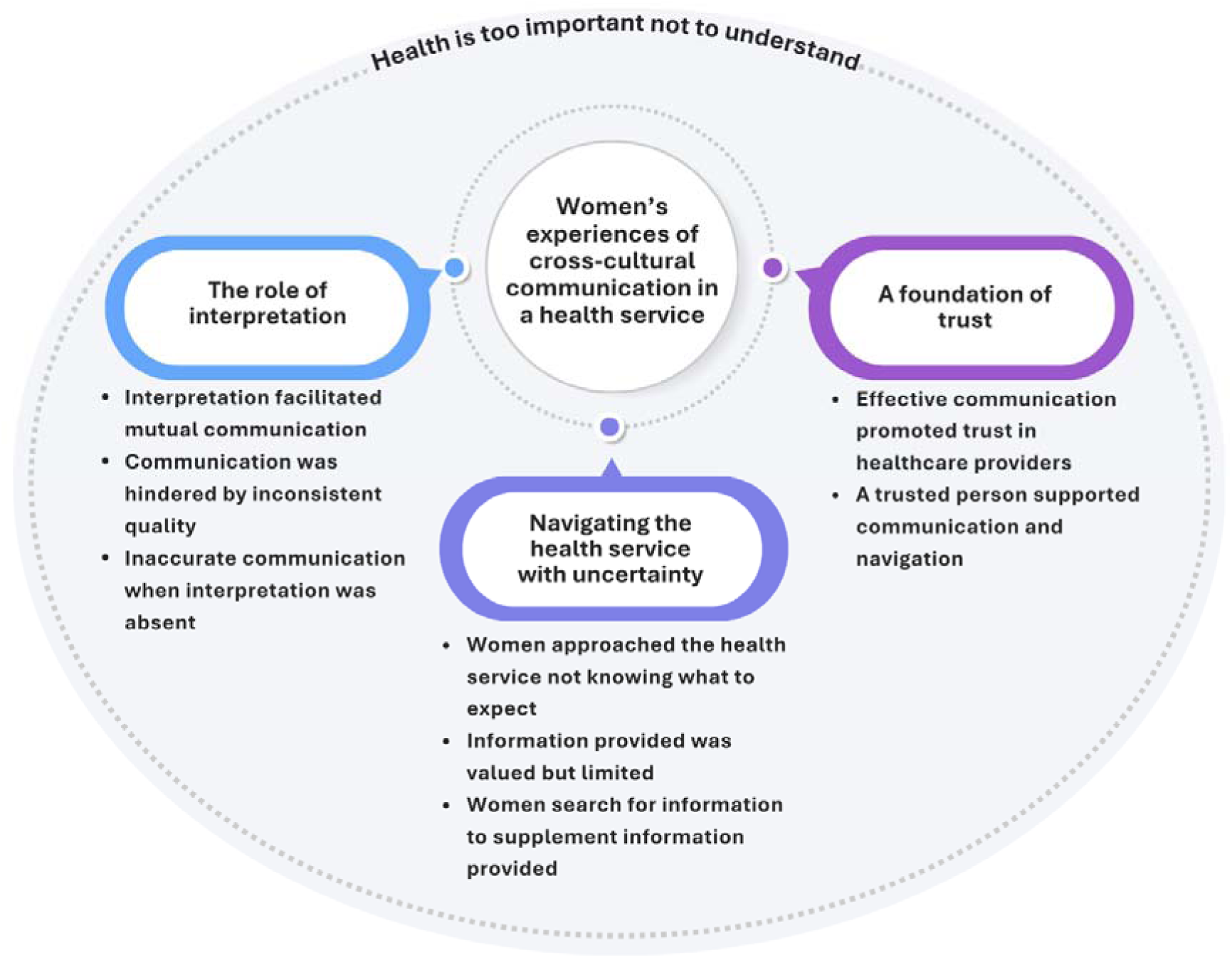
Thematic map of women’s cross-cultural communication experiences

Themes and subthemes are described below with supporting quotes presented in English in italics followed by a participant number or pseudonym. Superscript letters indicate language group: A for Arabic, B for Burmese, F for Farsi, M for Mandarin and V for Vietnamese. Following, the superscript letters M and G indicate if the participant accessed maternity or gynaecology care.

### Theme 1. The role of interpretation

Interpreters were arranged by the health service at specific points in women’s care, primarily during outpatient appointments or to gain consent for procedures. In person, an interpreter with a sound knowledge of medical terminology facilitated mutual communication between women and HCPs.

However, when interpreters appeared rushed, disengaged or unsure of health concepts, women were left without an understanding of their care. Women attempted to communicate around language barriers when interpreters were not made available to them, sometimes describing dissatisfaction with communication and/or their care.

#### Subtheme 1.1 Interpretation facilitated mutual communication

Women described a spectrum of English language skills, from not being able to communicate in English at all to being confident in everyday English but unfamiliar with medical terminology. In this context, women described how interpreters facilitated mutual communication between them and HCPs by helping them to understand what was being said, explain their concerns and ask questions.

> *“When in-person, if I’m confused with something or I don’t quite understand something, I can ask the interpreter at once and so the interpreter can explain it to me immediately. Therefore, it creates a connection between the interpreter and me” (P12^B,M^)*

They felt that communication was easier, more direct, efficient and friendly with an interpreter. Having an interpreter in-person was preferred by most because it allowed women, interpreters and HCPs to observe each other’s facial expressions and body language, improving the flow of communication. Effective interpretation enhanced women’s confidence in their understanding of their condition or proposed care, from which they could then make decisions about their health.

> *“My ability to read and write English is good, and I can do daily things easily by myself…I was not sure whether I would fully understand medical terms in English, and I needed to receive accurate information about the surgery that I needed, from my doctor” (P39^F,G^)*

#### Subtheme 1.2 Communication was hindered by inconsistent quality

Women recalled an array of barriers influencing effective interpretation. They reflected that not all interpreters appeared to have a sound knowledge of medical terminology or health concepts, resulting in misinterpretation and misunderstandings. One woman expressed concern that some interpreters appeared to add their own opinions to the information they interpreted on behalf of the HCP, “*The translation wasn’t very accurate. Also, when the midwife was explaining things to me and I had some questions, the interpreter would answer using her own experiences”* (Eva^M,M^). Others were concerned that some interpreters appeared rushed or disengaged. This was sometimes associated with prolonged wait times for outpatient appointments, exceeding the length of time the interpreter was booked for, *“…many times, interpreters had to leave when their booking time was over, even though my consultation with the medical professionals had not completed yet”* (P67^M,M^).

If the interpreter was unable to stay, interpreters were then engaged by telephone. However, using telephone interpreters was often associated with technical problems, long wait times and an apparent lack of staff knowledge about how to arrange them. Once connected, the interpretation itself was typically described as less accurate and communication was less direct.

Experiences of ineffective interpretation reduced women’s confidence in their understanding of their health and plan of care.

> *“…they asked me to sign the consent form for induction…A male interpreter [was connected by telephone], but the interpreting he did, I think he probably did not know how to interpret some of the professional information, especially the things about labour and delivery…I think I understood more listening to the English. His understanding was probably no better than mine” (Helen^M,M^)*

#### Subtheme 1.3 Inaccurate communication when interpretation was absent

When an interpreter was not present, women communicated within the limits of their English language skills and appreciated the efforts of HCPs to speak slowly to try to help them understand. Some asked a friend, family member or support worker present to assist them with communication. Others described using a translation or artificial intelligence app on their phone, although the translation could be inaccurate, *“…besides using my phone, I didn’t have any other methods [to interpret]…it wasn’t effective. It was quite inconvenient, as it took up a lot of the doctor’s time, and the translation wasn’t always accurate…”* (Kitty^M,M^). Another woman described trying to learn English medical terms in preparation for labour and birth, having been advised that an in-person interpreter would not be made available to her.

Without an interpreter, women described barriers to understanding what was being said by HCPs, to asking questions and to communicating their concerns. This caused worry and confusion, *“…there wasn’t any interpreter at the [emergency department]…I did not fully express myself on that occasion…Since the doctors here said it was all good, so I just, just listen and accept”* (Helen^M,M^). Some disengaged from care, *“since I didn’t understand what she was saying, I just ignored it”* (Khine^B,G^). Others sought alternate avenues to seek healthcare such as presenting multiple times to the emergency centre or contacting a trusted general practitioner (GP). Others consented to care without understanding what was happening.

> *“…[the midwife] said she needed to give me a shot of something, I just, I went a bit lost, didn’t really understand. I just replied. OK, OK. And [she] gave me the shot. Yeah, I did not know what she gave me” (Helen^M,M^)*

### Theme 2. Navigating the health service with uncertainty

Women described experiences of navigating the health service and their overall care characterised by uncertainty. Expectations of care and how it would be delivered were informed by previous healthcare experiences, norms in their country of origin and information sought online and from peers.

Uncertainty existed regarding their right to an interpreter, when these rights applied and what level of autonomy they had over the interaction. Women valued when information was communicated effectively about what to expect and treatment options; however, both information content and how it was communicated could be inadequate.

#### Subtheme 2.1 Women approached the health service not knowing what to expect

Many women described uncertainty about what to expect when engaging with the health service. This could include the purpose of each outpatient appointment, what type of HCP they would see and treatment options, *“…I just take my referral from my GP and to see the hospital…without any knowledge about what we will do next, yeah. So that’s make me a little bit worried…”* (Lily^M,M^). Women accessing gynaecological care specifically described uncertainty about how long to expect to wait for test results or surgery, and about the frequency and method of follow-up appointments. There was uncertainty about potential out-of-pocket costs, “*[I was]* c*oncerned that [an interpreter] might involve fees”* (P40^M,G^). Many women also expressed uncertainty about their right to an interpreter, when this right applied, and whether they could choose when they wanted an interpreter to be present and not to be present, or if they could request a specific interpreter.

Misconceptions were reported such as not having a right to an interpreter for telephone interactions with the health service or being ineligible to access continuity of midwifery care models. Repeated instances were described in which an interpreter was not made available including during telephone consultations, when presenting for emergency care and in labour and birth. Women found it difficult to advocate for themselves when they were unsure what was normal and what choices they had access to in Australia.

#### Subtheme 2.2 Information provided was valued but limited

Women described what information was provided to them about what to expect of their care, verbally and in writing. During appointments, women valued being told what may be discussed during their next appointment or when to expect to be contacted regarding test results and treatment. Outside of their appointments, information advising women about their next appointment or surgery was communicated in English by text message, email or letter.

> *“…it’s not good, because I have to give it to someone to read it to me…they know, for example, that this is an Arab who needs an interpreter [so] send them the letter…[in] Arabic so that they don’t miss the appointment…” (Dalal^A,G^)*

Typically, appointment notifications were restricted to a date, time and location. However, women wanted more practical information to allow them to plan appropriately, such as if an interpreter had been booked and a hospital map or directions, “*…the hospital could, for example, provide information on whom I need to see at each appointment and roughly how long the whole process will take”* (Mavis^M,M^).

Some women depended on a family member or support worker to explain information to them. Other women were confident in their English literacy skills and used an app to translate as needed. However, their comprehension could be hindered by tiredness and inaccuracies in web-based translations.

> *“…the hospital gave me a booklet. It contains details about how things develop week by week and which tests are needed…I just felt very tired when I was pregnant…since English, after all, is not my first language, I wasn’t much motivated to read it anyway.” (Eva^M,M^)*

#### Subtheme 2.3 Women searched for information to supplement information provided

Some women described speaking with other women from similar cultural backgrounds or searching language concordant online forums and social media apps to gauge what was normal with respect to their antenatal care or treatment for gynaecological conditions. However, they expressed uncertainty about the reliability and accuracy of information obtained outside of the health service. They valued being able to check this information with an HCP at the health service or a trusted GP, and information obtained from websites identified by HCPs as reliable, *“…when you just search on the Google, there were some people just do the chatting…but if there some…website that midwife give to us, or some book that midwife give to me…I can trust to follow it* (Lily^M,M^). Others were not confident or interested in searching for information online.

Online antenatal education classes offered by the health service were attended by some women; however, they were delivered in fast-paced English without an interpreter, making them difficult to understand. Some women described instead seeking language concordant information online such as Australian antenatal classes delivered online in Chinese, *“…both Chinese and English were provided [in the external class], and she…offered more information for me to understand how the delivery process unfolds in Australia, and what the environment was like in an Australian hospital*” (Helen^M,M^).

### Theme 3. A foundation of trust

Women approached the health service and HCPs with trust associated with HCPs’ perceived expertise. Many stated that trust was enhanced through good health outcomes, effective communication and acts of cultural safety. Continuity of care provider models were valued in both gynaecology and maternity care; however, this was rarely encountered. Having a trusted person such as an interpreter or support person by their side helped women navigate the health service with confidence, and interpreters could be viewed by women as having a support role. In cases where trust was established with interpreters, continuity of interpreter was valued, however this was also rarely encountered.

#### Subtheme 3.1 Effective communication promoted trust in healthcare providers

Women described approaching HCPs and the health service with trust which was associated with a perception of HCPs’ expertise. Trust between HCPs and women could be promoted through good health outcomes, effective communication, and a friendly and welcoming manner. Effective communication included effective interpretation, providing written information in women’s preferred language and HCPs who spoke women’s preferred language. Women also felt reassured when HCPs asked about their cultural needs, mental health and safety at home, *“…they also asked with care, they asked every time they met me, that if I was mentally well? Did I need any support? Did I feel safe?…At first, I felt a little strange. But then I felt more assured”* (Jade^V,M^).

Seeing the same HCP was valued when the woman had developed trust with that individual and continuity over multiple interactions allowed the HCP to learn how to communicate effectively with that woman, *“…if we can just meet the similar [same] midwife, that would be really helpful…he will just know how to communicate with me, or [if they need] to speak a little bit slow down so I quite understand it…”* (Lily^M,M^). However, continuity of care with HCPs was rarely encountered.

Some women described a loss of trust when communication broke down, interpreters were not provided, or their health needs were not met. One woman described a pre-existing distrust of HCPs due to a previous negative experience at a different hospital during which the poor care she had received had been blamed on her perceived lack of English language skills. She requested an interpreter at this health service as a measure to protect herself from HCPs and poor care, *“…I do not want something to happen to me and then [the healthcare providers] blame me for their inexperience and say that you do not understand the language”* (P61^F,M^).

*Subtheme 3.2 A trusted person supported communication and navigation* Having a trusted person to stand by their side throughout their care was valued by some women and desired by others, *“if I were to suggest an improvement, it would be having someone accompany me when I go for things like blood tests or check-ups in other departments, as sometimes I’m not familiar with the place”* (Kitty^M,M^). For some, female in-person interpreters could fulfil this role while for others, a support person such as a partner, family member or support worker accompanied them. Having someone by their side was a source of comfort and confidence for women. Support people were described as assisting women with communication alongside interpreters, and for some women, were preferred to a hospital-provided interpreter, *“Sometimes, I need to ask my support workers or caseworkers. They share their experiences and what they know with me, offering guidance and helping me”* (Khine^B,G^).

Women valued someone waiting with them for and between appointments, helping them to find different locations within the hospital and offering advice from their personal experiences about their health needs and care, and how to navigate the health service. For this reason, women valued having an interpreter provided in-person rather than by telephone. If an interpreter was viewed as professional and skilled, and they offered support and advice, trust could be established.

> *“…I really liked [the interpreter] because she…had two babies and she gave me lots of the experience what she said and lots of the details about that hospital, to tell tell me…what to do, and also let me [be] a little bit clear about if I meet this situation, what I maybe say. So she gave me lots of information that’s really helpful” (Lily^M,M^)*

Continuity of interpreter was valued in these instances, however, seeing the same interpreter throughout their care was rare.

## Discussion

This study offers insights into women’s experiences of cross-cultural communication in an Australian women’s health service. Quantitative findings reveal the diversity amongst women who use interpreter services and identify unmet communication and information needs. Qualitative findings highlight the key role of interpretation in facilitating effective communication, women’s uncertainty navigating the health service and the value of having someone trusted to support communication and navigation. Overall, the study highlights the need for a proactive service-wide approach to communication. Key systemic gaps are explored in the following discussion highlighting: inconsistent integration of interpretation services; lack of language concordant resources to promote informed decision-making, and a need for support with system navigation.

Our findings affirm the key role of professional interpretation in cross-cultural communication. However, while most participants reported an in-person interpreter was provided in their preferred language at their most recent visit to the health service, interpretation services were not provided consistently and interpretation quality varied. Inadequate interpretation services are a recurrent theme in maternity and broader health services, identified in both Australian (7, 11, 30, 31) and international literature(32–34). The value of professional interpretation to facilitate communication in hospital settings has been well established(14, 15), yet systemic barriers persist. One recent Australian mixed-methods study investigating barriers to interpreter access during the COVID-19 pandemic from HCP perspectives, identified barriers including limited availability of in-person interpreters, long wait times for telephone interpreters and an inflexible booking system(35).

These barriers reflect a broader systemic issue; when interpreters are engaged through external agency-based casual employment models rather than employed within health services, it limits health services’ abilities to improve the availability and accessibility of interpretation services.

Additionally, training and support for interpreters have been recommended to reduce variability in interpreter quality, particularly for clinical areas which require nuanced communication such as women’s reproductive health (36). Health service governance over support and training provided to external interpreters is limited in the current employment model. This approach may also limit the ability of health services to systematically collect data to monitor, evaluate and improve interpretation services(14) and diminishes recognition of interpreters as essential components of the healthcare system. Further research is recommended to explore sustainable models to better integrate professional interpreters into health services and to evaluate the impact of these models on clinical and economic outcomes.

The findings of our study may reflect a lack of easily accessible multilingual information resources and that interpretation services alone were oftentimes insufficient to meet the communication needs of women with LEP. Our study highlights a lack of language-concordant resources provided to participants to support verbal communication, promote informed decision-making and facilitate health service navigation. Less than half of the participants were offered written information in their preferred language. This aligns with the findings of a scoping review of international research into the patient experience among people with LEP which found a need for improvement in the provision of translated health resources(16). A qualitative Australian study investigating strategies to address health literacy issues in the provision of maternity care to culturally and linguistical diverse women(37) similarly identified a need for improved provision of culturally and linguistically appropriate eHealth resources as adjuncts to verbal communication. The authors specifically recommended a hospital-endorsed multilingual health information app.

When exploring pathways to improve availability of multilingual resources provided to women with LEP, translating materials designed for English-proficient audiences can limit their relevance and acceptability(38). Our findings show that women with LEP may have specific information needs associated with navigating an unfamiliar health system which may not be met through translation alone. The importance of participatory approaches in the design and implementation of health information resources to ensure they are responsive to the needs of culturally and linguistically diverse populations has been emphasised in maternity and health promotion settings(37, 39).

Adopting a co-design approach more broadly within hospital settings is warranted to ensure women have access to language-concordant resources responsive to their needs.

Our study participants described uncertainty navigating the hospital system exacerbated by English-language hospital correspondence lacking information content to support service navigation. In their scoping review of the patient experience, Yeheskel and Rawal(16) similarly reported appointment letters and forms issued in English only, associating this English language communication with poor appointment attendance. A content analysis of acute care hospital websites located in one state in the United States(40) further found that only 10.8% provided translated content and while 81.7% of websites identified the availability of interpretation services, this information could only be accessed after navigating through multiple English language webpages. These findings demonstrate the challenges that people with LEP can face to supplementing inadequate hospital communication by searching online. The study authors recommended that hospitals improve their online communication strategies to meet the needs of linguistically diverse communities(40). Our findings highlight the need for proactive language-concordant communication strategies beyond clinical interactions to effectively support service navigation by women with LEP.

Our study highlights the value women placed on having someone trusted to support them with service navigation, often referencing advice and support provided by an interpreter. Service navigation however, is beyond the scope of professional interpreters in Australia(41). The Australian Institute of Interpreters and Translators Code of Ethics and Code of Conduct(41) outlines the role of interpreters as facilitators of communication through message transfer, precluding their involvement in advocacy, guidance or advice.

Despite this, interview participants described instances of interpreters acting outside this scope to provide assistance and advice, with some appreciating this and others not. This suggests a gap in advocacy and support for service navigation. Previous research has explored language-concordant health navigator or cultural broker roles in Australia(42) and internationally(43).

DiMeo et al(43), in their critical review of cultural broker roles in pregnancy care, emphasise the importance of role clarity and a supportive ecosystem for the role to be successful. Our findings suggest there is a role for navigators, however, further research is needed into the sustainable implementation and impact of language-concordant health navigator roles within hospital settings.

### Strengths and limitations

This study provides much needed data on consumer perspectives of cross-cultural communication. Unlike previous research that identified HCP skills and training as an area for improvement(7), participants in this study reflected little on this aspect of their care. This may be because the study was conducted by the health service that they had received or were continuing to receive care through. Although the research team were not involved in the participants’ clinical care and participants were informed that their participation would not impact their care, this power dynamic may have affected the study findings. The small sample size may also be considered a limitation of this study; however, the use of a mixed methods design is a strength(17). Triangulation of quantitative and qualitative data offered insights into aspects of women’s experiences not otherwise captured. Throughout the design and conduct of this study, an emphasis was placed on cultural responsiveness and language concordance, exemplified through the crucial role of bicultural bilingual research assistants in tool design, data collection and analysis. It is possible however that nuances in women’s experiences were missed due to language or cultural discordance.

## Conclusion

In an increasingly multicultural world, cross-cultural communication requires a proactive whole of health service approach to meet the communication and service navigation needs of women with LEP. This study shows that women’s experiences of communication extend beyond clinical interactions, however support for communication largely did not. Within clinical interactions, provision and quality of interpretation services were variable. Planning and resourcing to support effective cross-cultural communication, particularly with respect to the integration of interpretation services and language-concordant support for service navigation, must be key considerations in the delivery of healthcare for service and clinical area leads, and policymakers.

## Declarations and ethics statements

### Ethical approval

This study received approval from the Western Australian Health Service Human Rights Ethics Committee (HREC) (approval number RGS0000005315) and Curtin University HREC (approval number 2022-0508).

### Informed consent from participants

Informed consent was obtained from all study participants. A participant information sheet was made available in the language spoken by their interpreter and English, and participants were provided an opportunity to ask questions of a female language concordant research assistant by phone.

Survey participants confirmed their eligibility to participate and provided consent with a ‘hurdle’ survey item prior to commencing the survey. A consent form was read out to interview participants in their preferred language by an investigator. Interview participants provided verbal consent. The investigator then signed the consent form as having witnessed the participant’s verbal consent.

## Data availability statement

The data that support the findings of this study are available on reasonable request from the corresponding author, GG. The data are not publicly available due to ethical restrictions.

## Author contributions

Conceptualisation: GG, NW, JW, ZB. Funding acquisition: GG, NW, JW, ZB. Methodology: GG, NW, NJ, JW, ZB. Project administration: GG. Investigation: GG, SG, YSL, MMZ, MM, SN, TKAN. Formal analysis: GG, SG, YSL, MM, TKAN. Visualisation: GG. Writing – Original draft: GG. Writing – Review and editing: SG, YSL, MMZ, MM, SN, TKAN, NW, NJ, JW, ZB. Supervision: NW, NJ, ZB. All authors have read, critically revised, and approved the final manuscript.

## Data Availability

All data produced in the present study are available upon reasonable request to the authors.

## Acknowledgements

The authors acknowledge the contributions of the research assistants who made this research possible. We are sincerely grateful for the women who generously shared their experiences of cross-cultural communication at the health service towards improving the experiences of women in future. Thank you also to the clinicians and researchers who supported the conduct of this study.

