## Supplementary material for "Cross-cultural communication in a women’s health service: A mixed-methods evaluation"

**Cross-Cultural Communication Project Survey Tool**

*The English language survey tool presented below was translated for online and telephone data collection into Arabic, Burmese, Farsi, Mandarin and Vietnamese.*

First, please confirm that you have read the Participant Information Form or had the Participation Form read to you, that you received maternity or gynaecology care at [hospital site 1] or [hospital site 2] and consent to participating in this survey. You can find a copy of the Participant Information Form at this link.

- Yes, I have read the Participant Information Form and I understand the purposes, procedures and risks of the research described in the project. I understand that I am free to withdraw up until submission of my survey responses, and that withdrawing will not affect my future health care. I freely consent to participate in this research project as described.
- Yes, I received maternity or gynaecology care at [hospital site 1] and/or [hospital site 2].

These next questions are some information about you.

1. What is your age? _____________________
2. What did you attend [the health service] for?

- Urological gynaecology (problems with your bladder)
- General gynaecology (problems with your uterus/ovaries/vagina that are not related to cancer)
- Gynaecological cancer
- Antenatal care (clinics)
- Pregnancy complications (e.g. Maternal Foetal Assessment Unit/Emergency Centre)
- Labour and birth
- Postnatal care
- Pregnancy loss or complications (less than 20 weeks)
- Pregnancy loss (more than 20 weeks)
- Other: ________________

3. What ethnicity do you identify as? ______________
4. What is your country of birth? ________________
5. How many years have you lived in Australia? _________________
6. What is your religion? ________________________
7. What is the primary language you speak? ______________
8. How well can you read in your primary language?

- Not at all
- Not well
- Well
- Very well

9. How well can you write in your primary language?

- Not at all
- Not well
- Well
- Very well

10. How well can you read in English?

- Not at all
- Not well
- Well
- Very well

11. How well can you write in English?

- Not at all
- Not well
- Well
- Very well

12. In your most recent visit, what language did the interpreter speak? _____________
13. Was this your preferred language?

- Yes
- No

14. Did you ask for an interpreter or was an interpreter offered by staff?

- Asked *(Branching logic to Q15)*
- Offered *(Branching logic to Q16)*
- Not sure *(Branching logic to Q16)*

15. Please say why you asked for an interpreter
16. Did you feel confident to ask for an interpreter?

- Yes
- No

17. Please say why you did or did not feel confident
18. What ways have you communicated with an interpreter at this hospital?

- In-person
- Telephone
- Video conference/Telehealth
- iPAD app

19. Please rank interpreter type in order of preference:

- In-person
- Telephone
- Video conference/Telehealth
- iPAD app

20. Why did you rank [*selection*] as your first preference?
21. Did you feel comfortable communicating with the healthcare provider (midwife, nurse, doctor, physiotherapist etc) through the interpreter?

- Yes
- No

22. What makes you feel comfortable when communicating with a healthcare provider?

- Female healthcare provider
- Female interpreter
- The same healthcare provider I’ve seen before
- The same interpreter I’ve seen before
- Friendly healthcare provider
- Friendly interpreter
- Family member, friend or support worker present
- Other: __________________________________________

23. Were you offered written information in English or your primary language?

- Yes, in English
- Yes, in [language]
- No
- Not sure

24. Would you like to be offered written information?

- Yes, in English
- Yes, in [language]
- No

25. Do you feel confident to ask for written information?

- Yes
- No

26. Are you aware of the [health service] Library as a place to access written health in information in [language]?

- Yes, I already use the library
- Yes but I don’t use the library
- No

27. Do you use technology to communicate about or find information about your [pregnancy/condition]?

- Yes, I use a phone app: _________
- Yes, I use a computer
- No

28. Do you use the internet to find information about your [pregnancy/condition]?

- Yes, I already do *(Branching logic to Q28a)*
- No but I would like to *(Branching logic to Q28b)*
- No, I don’t want to *(Branching logic to Q29)*

28a.	If you use the internet, how do you make sense of information that is not in [*primary* language]?

- Use a translation app or website (e.g. Google Translate)
- Ask a friend/family member/child
- Look at videos or pictures
- Only access information in my primary language
- Only access information from my country of origin
- Other: _______________________

28b.	If you would like to use the internet to find information about your [pregnancy/condition], what is stopping you?

- Don’t have technology skills
- Don’t know where to look
- Not sure what are reliable websites
- Find it difficult to translate information
- Don’t have access to the internet
- Too busy
- Other: _______________________

29. This is the end of the survey. Once I submit your response, you will no longer be able to withdraw. Do you consent for your responses to be submitted?

- Yes

**Cross-Cultural Communication Project Interview Guide**

*The English language interview guide was translated into Arabic, Burmese, Farsi, Mandarin and Vietnamese.*

**Demographic questions**

1. What is your age? _____________________
2. What did you attend [the health service] for?

- Urological gynaecology (problems with your bladder)
- General gynaecology (problems with your uterus/ovaries/vagina that are not related to cancer)
- Gynaecological cancer
- Antenatal care (clinics)
- Pregnancy complications (e.g. Maternal Foetal Assessment Unit/Emergency Centre)
- Labour and birth
- Postnatal care
- Pregnancy loss or complications (less than 20 weeks)
- Pregnancy loss (more than 20 weeks)
- Other: ___________________

3. What ethnicity do you identify as? ______________
4. What is your country of birth? ________________
5. How many years have you lived in Australia? _________________
6. What is your religion? ________________________
7. What is the primary language you speak? ______________

**Interview Questions**

1. What helped you to communicate with the healthcare provider at the hospital?
2. What made it difficult to communicate with the healthcare provider at the hospital?
3. How can healthcare providers at the hospital make it easier for you to communicate with and understand them?
4. What made you feel confident and safe to ask questions and communicate at the hospital?
5. What would make you feel safer and more confident at the hospital?
6. How would you like technology or the internet to help you communicate or find information about your [pregnancy/condition]?
7. What would make it easier to use technology or the internet to communicate or find information about your [pregnancy/condition]?
8. Is there anything else you would like us to know about your experiences?

**Additional prompts**

- Can you say more about that?
- Can you give me an example?
- What was that like for you?
- You said earlier….
- How do you think other women….?
- Is that your experience too?
- We have been talking about when you were…, can you tell me…?

**Positionality Statements**

YSL is a Burmese international student who has worked on research with vulnerable communities, including those in culturally and linguistically diverse populations. As a young woman passionate about mental health, her lived experiences navigating healthcare systems have shaped her understanding of the barriers faced by Burmese women, particularly refugees, in accessing healthcare in Australia.

MM is an Iranian PhD student in Australia, researching privacy and security in healthcare data. Her background as a Farsi-speaking immigrant facilitated communication with women, encouraging them to share their experiences. It also informed her perspective on how this research could benefit others by enhancing cross-cultural communication in hospitals. Her focus on ethical data handling likely deepened her understanding of women’s concerns about having a friendly and trusted interpreter or healthcare provider with whom they can confidently share their information.

TKAN is an interpreter who undertakes interpreting at various hospitals including the study setting. She joined the project as she is committed to improving community healthcare quality in general and communication in this area in particular. Working as an interpreter as her daily work gives her the opportunity to understand some of the challenges facing the health care system, especially those experienced by the overloading at many public hospitals. As a migrant to Australia, she understands the difficulties of those in her community who have language barriers, especially for women who need treatment for their women’s health issues in a hospital setting.

Originally from Iraq and a native Arabic speaker, SG was instrumental in developing Arabic-language research materials, and conducted all interviews with Arabic-speaking women, creating a respectful and culturally sensitive environment that encouraged open dialogue. This was facilitated by her insider stance within the Arab community.

GG, who led the project design and conduct, held an outsider stance regarding the experiences of women with low English proficiency and an insider stance as a clinician with nursing experience at one of the study sites. This insider knowledge of the health service helped her to explore women’s experiences during data collection. She maintained an awareness of this positionality through structured team reflexivity discussions.

Another research team member brought her experience as a researcher and data analyst to the project. As a native Mandarin speaker from China, she had firsthand cross-cultural communication experiences at the health service in Australia. She assisted with the design of recruitment materials, conducted all interviews with Mandarin-speaking participants and conducted data analysis. As an insider, she promoted participant recruitment, streamlining the process and ensuring a comfortable environment for women to share their experiences during phone interviews.
